# Efficient and Safe Outpatient Shoulder Arthroplasty Using Isolated Regional Anesthesia: A Retrospective Cohort Study

**DOI:** 10.1101/2025.09.15.25335825

**Authors:** Ibrahim Al-Marei, Justin Jewell, Jesse Ng, David Furgiuele, Germaine Cuff, Michael Reynolds

**Author notes:** Corresponding Author: Michael Reynolds, MD, Department of Anesthesiology, Perioperative Care, and Pain Medicine, NYU Langone Health, 550 1st Avenue T-530, New York, NY 10016. Funding: None.

## Abstract

**Background:** Regional anesthesia as a sole modality for total shoulder arthroplasty (TSA) remains rare in the U.S., typically <2% of cases. Benefits—reduced opioid use, shorter recovery, fewer airway/general anesthesia complications—are well-known, but real-world ambulatory data remain scarce. We aimed to evaluate efficacy and safety of interscalene block with sedation as the primary anesthesia modality in an outpatient TSA population.

**Methods:** This retrospective study reviewed 330 adult elective primary TSAs from January 2018 to December 2024 at a tertiary academic center. Interscalene block using 0.5% long acting local anesthetic with sedation was used in all. Primary outcomes included: ambulatory discharge success, conversion to general anesthesia. Secondary outcomes included: PACU duration, time from anesthesia start to ready, complications, 30-day readmission. Data were compared to national benchmarks.

**Results:** Conversion to general anesthesia was required in 4 of 330 cases (1.2%). Ambulatory discharge success rate, was achieved in 95.9% of patients. Mean PACU stay was 57.4 ± 14.8 minutes. Anesthesia start-to-ready time was 12.1 ± 3.6 minutes. No major anesthesia-related complications occurred except for transient hemidiaphragmatic paresis seen in 1.2% of patients. Readmission rate was 2.7%, none anesthesia-related.

**Conclusions:** Regional with sedation anesthesia for TSA demonstrated a high ambulatory discharge success rate with minimal complications. This model is feasible for high-throughput ambulatory centers.

## INTRODUCTION

Total shoulder arthroplasty (TSA) is rapidly evolving toward outpatient care, driven by cost savings, enhanced recovery protocols, and improved surgical techniques^1,2^. However, anesthetic approaches remain variable. General anesthesia (GA), often supplemented by regional nerve blocks, remains the most frequently utilized anesthetic. Recent NSQIP data from 2005–2018 show that regional anesthesia (RA) as the primary anesthetic accounts for just 1.6% of TSA cases nationwide^3,4^, with an overall trend of regional anesthesia as the primary anesthetic decreasing over time^3^.

Despite its lower utilization rate, regional anesthesia with or without sedation offers several advantages: elimination of airway manipulation, reduction of postoperative nausea and sedation, lower opioid use, and decreased PACU length of stay^5–7^. Still, real-world data demonstrating the safety and efficacy of RA-only models for TSA, especially in ambulatory settings, are sparse.^7^

Our institution developed a practice model leveraging single-shot interscalene block with sedation for outpatient TSA. We hypothesized this technique would support a high ambulatory discharge success rate with low complication rates. This study describes our experience using this regional-only model and compares outcomes to both national data and historical GA-era benchmarks.

## METHODS

This was a single-center retrospective cohort study at a tertiary academic medical center. Adult patients undergoing elective primary TSA between January 2018 and December 2024, aged 18-80 years of age, and ASA class 1-3 were identified through EMR query. IRB approval was obtained (protocol #i24-01901).

All patients received intraoperative ultrasound-guided interscalene block using up to 20 mL of 0.5% long-acting local anesthetic, typically bupivacaine. Sedation with midazolam, fentanyl, and/or ketamine with propofol with or without dexmedetomidine infusion was administered based on anesthesiologist preference. Conversion to GA remained standard backup if patients experienced block failure, respiratory compromise, or intolerance. Postoperatively, patients were managed with a multi-modal enhanced recovery pain regimen protocol.

Primary outcomes: Ambulatory discharge success, defined as discharge within 23 hours of surgery and conversion from RA to GA. Secondary outcomes: PACU time, time from anesthesia start to ready-for-incision (ART), 30-day readmission, block-related complications. Exploratory: ASA classification effects and time-of-day effects.

Descriptive statistics were calculated. Internal comparisons were made to a GA cohort. National data were referenced from NSQIP, UCSF, and peer-reviewed publications.

## RESULTS

A total of 330 regional-only TSA cases met inclusion criteria. The cohort had a mean age of 61 years and a mean BMI of 30.12. The sex distribution was 56.7% female and 43.3% male. ASA classification was as follows: ASA I in 6 patients, ASA II in 235 patients, and ASA III in 89 patients (Table 1).

**Table 1.** Patient Demographics and ASA Class. Summary of baseline demographic characteristics and ASA physical status classification for the 330 patients included in the regional-only cohort.

| Characteristic | Value |
| --- | --- |
| Total Patients | 330 |
| Mean BMI | 30.12 |
| Sex, Female | 56.7% |
| Sex, Male | 43.3% |
| ASA Class I / II / III | 6 / 235 / 89 |

A conversion rate of 1.2% to general anesthesia and an ambulatory discharge success rate, defined as discharge the same day or within 23-hours, of 95.9% was observed. Hemidiaphragmatic paresis (HDP) rate (1.2%) was consistent with literature supporting volume-related risk.

Though our study was not powered to perform a definitive statistical comparison between anesthesia types, internal analysis of anesthesia start-to-ready (ART), OR, and PACU times across our GA and regional-only cases reveals meaningful trends. ART was 12.1 minutes for regional-only cases compared to 16.1 minutes for GA ± RA (p = 0.068). OR time was significantly shorter in the RA group (168 vs. 195 min, p = 0.018), while PACU time was also reduced (57.6 vs. 86.9 min, p = 0.109). These findings suggest efficiency gains with regional-only anesthesia, despite not reaching statistical significance for all outcomes.

## DISCUSSION

In this single-center case series, we report a regional with sedation anesthetic approach for total shoulder arthroplasty with a 95.9% ambulatory discharge success rate and a 1.2% conversion to general anesthesia, none of which occurred for emergent airway or hemodynamic concerns. This model remains rare in U.S. practice, accounting for just 0.83% of TSAs between 2010 and 2015 according to national registry data^3^, yet demonstrates excellent outcomes when implemented within a structured and experienced protocol.

Ambulatory discharge success remained high across ASA classes, including 95.5% in ASA III patients (Figure 1), and was slightly lower in revision cases compared to primary cases (Table 2), consistent with the expectation that surgical complexity may influence postoperative disposition.

**Table 2.**
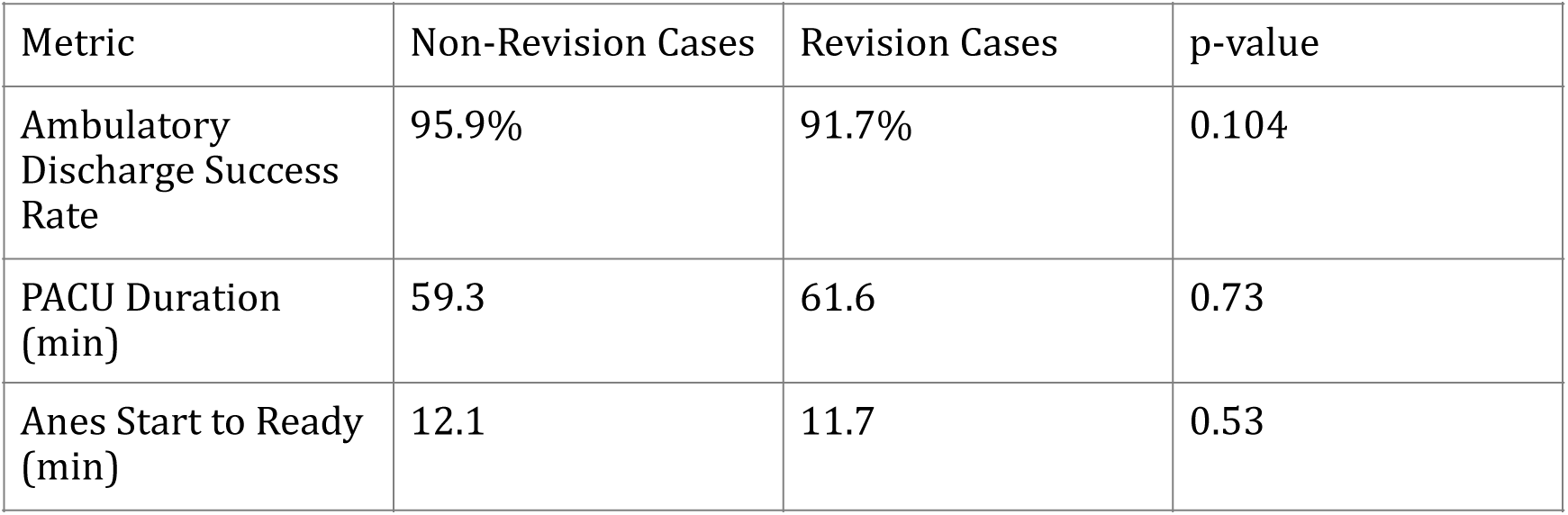
Ambulatory Metrics of Primary versus Revision Total Shoulder Arthroplasty. Comparison of ambulatory discharge success rate, post-anesthesia care unit (PACU) duration, and anesthesia start-to-ready time between primary and revision total shoulder arthroplasty cases. No statistically significant differences were observed across the reported metrics.

**Figure 1.**
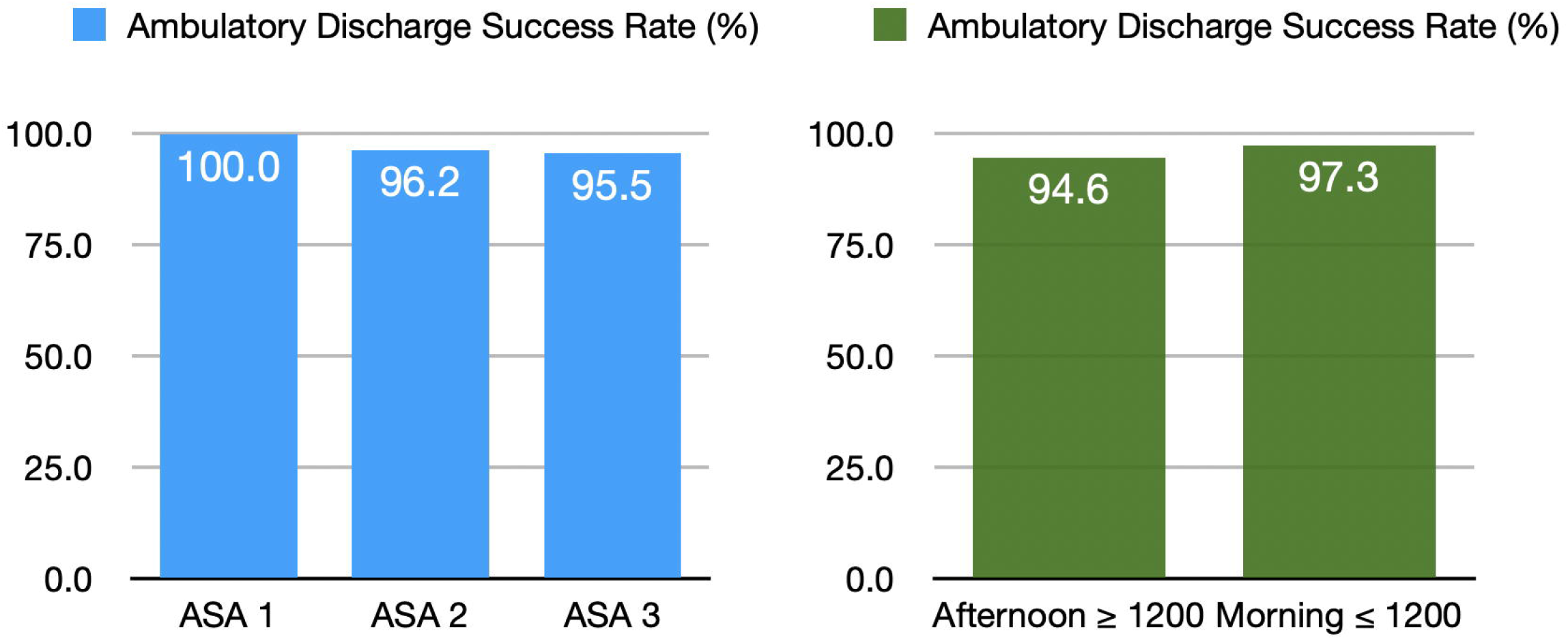
Ambulatory Discharge Success Rates by ASA Class and Case Start Time. Discharge success rate (%) by patient ASA classification (left) and case start time (right). Discharge success was defined as home discharge within 23 hours without unplanned admission.

A modest decline in discharge success for afternoon cases (94.6%) relative to morning cases (97.3%) (Figure 1) may reflect operational or recovery-related constraints rather than patient-specific factors.

In our cohort, the regional-only approach was associated with low rates of complications, unplanned admissions, and readmissions (Table 3).

**Table 3.** Key Outcome Comparison Between Our Regional-Only TSA Model and National Data. National benchmark values derived from previously published datasets for general anesthesia (GA)-only cases, GA with regional anesthesia (RA), and RA-only cohorts. Reference ranges are reported where available. Sources: Gabriel et al.^3^, Bixby et al.^4^, Memtsoudis et al.^9^

| Metric | Our Regional-Only Model | National - GA Only | National - GA + RA | National - RA Only |
| --- | --- | --- | --- | --- |
| Mean OR Time (min) | 168 | 105.6–139.0 | Similar to GA | Not well defined (rare use) |
| Mean PACU Time (min) | 57.4 | Not reported | Not reported | Not reported |
| Conversion to GA Rate | 1.2% | Not reported | Not typically reported | Varies widely |
| Unplanned Admission Rate | 3.9% | ~6–8% | ~5–6% | <5% |
| 30-Day Readmission Rate | 2.7% | 2.5% | 1.8% | <1.5% |
| Pulmonary Complication Rate | 1.2% (3/4 due to HDP) | 1.4% (baseline) | Reduced vs GA alone | Lowest rate |
| Ambulatory Discharge Success Rate | 95.9% | 85–91% | 90–93% | Highest (95–100%) |

These outcomes are consistent with national database analyses. Memtsoudis et al. reported that regional anesthesia alone was associated with the lowest rates of pulmonary complications and adverse perioperative outcomes in orthopedic surgery patients, including those undergoing total shoulder arthroplasty.^9^ Stundner et al. found that the addition of peripheral nerve blocks to shoulder arthroplasty was associated with lower complication rates and shorter hospital stays.^10^ These findings support the safety and effectiveness of regional anesthesia as a primary anesthetic in shoulder arthroplasty, particularly in the ambulatory setting.

In addition to reducing complications, the regional-only approach may enhance overall perioperative efficiency by facilitating faster recovery milestones. Our cohort demonstrated a mean PACU time of just 57.4 minutes, substantially shorter than national benchmarks that often exceed two hours. These findings suggest that this technique may contribute meaningfully to institutional goals of same-day discharge and perioperative efficiency, especially relevant for systems seeking to reduce OR and PACU utilization while maintaining safety. In a large managed-care study, Yian et al. reported a 22% cost reduction for outpatient TSA compared to inpatient procedures, largely driven by lower facility and implant costs.^11^ Grobet et al. similarly reported that shorter hospital stays and outpatient surgical settings were associated with improved cost-effectiveness, based on real-world prospective economic modeling.^12^

Hypoxia and pain were the two most common reasons for delayed discharge. Hemidiaphragmatic paralysis, responsible for 75% of hypoxia-related events, was the most frequent anesthetic complication (0.9%). This aligns with the known dose-dependent nature of interscalene blocks,^13^ and the close anatomic relationship between the brachial plexus and the phrenic nerve at the level of the interscalene groove. Tran et al. further highlight that even small changes in needle tip position and local anesthetic volume may meaningfully impact phrenic nerve spread, reinforcing the value of precision-guided, anatomically informed approaches.^14^ In a comprehensive review published in Anesthesiology, El-Boghdadly et al. reported that the incidence of HDP approaches 100% with 20 mL of local anesthetic, but may decrease to 30–40% with volumes of 5 mL or less, emphasizing the importance of volume selection and needle positioning.^15^ A recent meta-analysis by Oliver-Fornies et al. confirmed that these diaphragm-sparing techniques can significantly reduce the incidence of complete hemidiaphragmatic paralysis by 65%.^16^ In patients at risk of pulmonary compromise, lower-volume strategies and alternative approaches such as the superior trunk, suprascapular, or costoclavicular blocks may reduce the risk of phrenic involvement while maintaining surgical anesthesia.^17–22^

Ten patients (3.0%) experienced pain-related delays in discharge. For patients identified preoperatively as being at risk for poorly controlled perioperative pain, such as those who are younger, female, or report significant preoperative pain,^23^ continuous peripheral nerve catheters can be selectively employed in practice.^16,24^ A targeted use of catheter-based analgesia allows for extended block duration without committing all patients to catheter placement, management, and follow-up and can enhance individualized care within a regional anesthesia framework.^6^

Concerns around airway access in the beach-chair position are often cited as a rationale for defaulting to general anesthesia. However, published conversion rates from sedation to general anesthesia in this setting range from 2–6%,^25^ and our observed rate of 1.2%— with no emergent events—compares favorably. Moreover, spontaneous ventilation with light sedation may confer hemodynamic stability advantages over general anesthesia, particularly in the beach-chair position where cerebral perfusion can be precarious.

Prospective monitoring has demonstrated diminished cerebral autoregulation and lower cerebral oxygenation in this position,^26^ but importantly, without associated postoperative cognitive or biomarker evidence of neurologic injury. With appropriate patient selection, team familiarity, and defined escalation pathways, regional anesthesia with sedation can be delivered safely.

For anesthesiologists seeking to incorporate a regional with sedation model into their TSA practice, surgeon collaboration is essential. Successful implementation depends on shared expectations regarding surgical workflow, airway management readiness, and discharge planning.^27^ Importantly, orthopedic literature also supports the safety and efficacy of this approach. A retrospective propensity-matched study from the NYU Department of Orthopedic Surgery found that patients undergoing TSA under isolated regional anesthesia had lower in-hospital complication and readmission rates compared to those receiving general anesthesia.^8^ These findings are consistent with national-level analyses. Vallabhaneni et al. found that patients receiving regional anesthesia alone for TSA had significantly lower pulmonary complication rates, reduced length of stay, and lower hospitalization costs compared to those receiving general anesthesia.^28^ Herrick et al. similarly found that regional anesthesia for TSA was associated with lower ICU admission rates, reduced 30-day complications, and lower overall costs.^7^

In our cohort of over 300 patients undergoing total shoulder arthroplasty with a regional-only approach, the incidence of serious anesthesia-related complications was remarkably low. There were no cases of local anesthetic systemic toxicity, recurrent laryngeal nerve palsy, or prolonged neurologic deficit. The only observed anesthetic complication was hemidiaphragmatic paralysis, which was transient in all cases. When performed with strict adherence to aseptic technique and established dosing guidelines, regional anesthesia offers a favorable safety profile even in high-volume regional anesthesia practices.^29^

Although alternatives such as local infiltration analgesia (LIA) have been explored in total shoulder arthroplasty, studies suggest inferior analgesic efficacy. In a prospective cohort study, LIA resulted in significantly higher opioid consumption and worse pain scores in the first 24 hours postoperatively compared to interscalene block, despite similar discharge times and complication rates.^30^ This reinforces the continued value of regional anesthesia in optimizing early recovery and opioid stewardship.

Surgeon buy-in and consistent OR workflows were critical to success. Recent orthopedic anesthesia reviews have highlighted regional anesthesia as central to enhanced recovery protocols, citing benefits in cognitive preservation, hemodynamic stability, and discharge readiness.^4^ Despite strong safety and efficiency metrics associated with regional anesthesia, national utilization remains low. Cultural and logistical barriers likely play a role. In a survey of 468 orthopedic surgeons, Oldman et al. found that although 84% of surgeons who directed anesthetic care favored regional anesthesia, many cited perceived delays and unreliability as key limitations.^31^ Strategies such as dedicated block rooms, protocol-driven workflows, and enhanced surgical buy-in may be essential to improving adoption. As outpatient total joint arthroplasty expands, regional anesthesia with sedation offers a pathway toward higher-value care - particularly in high-volume ambulatory settings where efficiency, safety, and patient-centeredness must align.

Limitations include retrospective design, single-center setting, and lack of patient-reported outcomes. However, the consistency of outcomes across a large number of cases, including ASA III and revision procedures, supports the potential for replication at other institutions with similar patient populations and surgical workflows. Future multi-center prospective studies should assess reproducibility, economic impact, and long-term satisfaction.

These findings suggest that, in appropriately selected patients, regional anesthesia alone can provide safe and efficient anesthetic care for ambulatory total shoulder arthroplasty. When supported by multidisciplinary coordination, institutional buy-in, and protocol-driven patient selection, this model offers a practical and scalable alternative to general anesthesia - particularly in high-volume outpatient settings.

## Data Availability

All data produced in the present study are available upon reasonable request to the authors

## ACKNOWLEDGEMENTS

The authors wish to thank the NYU Langone Orthopedic Hospital Quality and Safety team for their assistance with data extraction and chart review.

